# Factor Structure of the Postpartum Bonding Questionnaire in a US-Based Cohort of Mothers

**DOI:** 10.1101/2023.04.09.23288334

**Authors:** Andréane Lavallée, Jennifer M. Warmingham, Mark A. Reimers, Margaret H. Kyle, Judy Austin, Seonjoo Lee, Tyson Barker, Maha Hussain, Sharon Ettinger, Dani Dumitriu

## Abstract

As research efforts in the field of pediatrics are oriented toward a better understanding of the synergistic relationship between different facets of early relational health (ERH) and child development and wellbeing, it is essential to focus on the quality of research instruments available for measuring different components of ERH. This study investigates the measurement characteristics of a widely used parent/caregiver-reported measure of bonding, the Postpartum Bonding Questionnaire (PBQ), in a US-based sample (n=610) of English-speaking biological mothers who completed the PBQ at 4 months postpartum. To evaluate the factor structure of the PBQ, confirmatory and exploratory statistical techniques were employed. The current study failed to replicate the PBQ’s original 4-factor structure. Exploratory factor analysis results supported the creation of a 14-item abbreviated measure, the PBQ-14. The PBQ-14 showed evidence of good psychometric properties, including high internal consistency (ω=.87) and correlation with depression (*r*=.44, p<.001) assessed using the Patient Health Questionnaire (PHQ-9), as would be expected. The new unidimensional PBQ-14 is suitable for use in the US as a measure of general postnatal parent/caregiver-to-infant bonding.

## Introduction

Promoting parent/caregiver-child early relational health (ERH) is now recognized as a priority by American and Canadian pediatric societies.^1,2^ Although decades of research supports the centrality of relationships for child development,^1,3-13^ we still lack a global understanding of ERH phenotypes and their respective role in predicting and explaining ‘for who, when and how’ different child-oriented life-course outcomes develop. To that effect, valid and reliable measures that accurately capture different components of ERH, such as parent/caregiver-reported bonding, are critical.

The Postpartum Bonding Questionnaire (PBQ) is the most widely used and researched parent/caregiver-reported screening instrument for bonding.^14,15^ Albeit often conflated,^15,16^ ‘bonding’ and ‘attachment’ describe components of parent/caregiver-child ERH that are, at the core, fundamentally different. Owing its origin to Klaus and Kenell,^17^ four decades of ERH research has led to a degree of agreement that bonding is a parent/caregiver-driven concept describing the parent/caregiver-to-infant emotional tie, from birth and beyond.^16,18,19^ In contrast, attachment is a child-centered component of ERH, manifested by secure, organized, insecure and/or disorganized behavioral patterns when faced with uncomfortable or distressing situations.^20^

The PBQ was originally developed in English and validated in the United Kingdom.^21^ Since first published in 2001,^21^ the PBQ has been used for research and clinical purposes across the world, and is translated and validated in at least nine languages, i.e., Italian,^22^ Portuguese,^23,24^ Japanese,^25-27^ French,^28^ Spanish,^29^ Tamil,^30^ Dutch,^31^ Bengali,^32^ and German.^33^ Despite its widespread adoption and otherwise acceptable psychometric properties,^15^ there remains uncertainty over the factor structure (i.e., empirical item groupings to inform subscale scoring).^21^ The initial factor structure from the 25-item measure originally yielded four factors^21,34^ defining general bonding disorders, severe mother-infant relationship disorders, infant-focused anxiety, and risk of abuse. Although the measure is often scored using the four factors listed above, this structure has never been confirmed or replicated.^15^ Other psychometric studies have suggested different factorial structures (see Table 1), varying from 1-factor^23,25,28,31,33^ to 4-factor structures,^29,32^ and retaining from eight^23^ to all 25 items.^26,29^

**Table 1.**
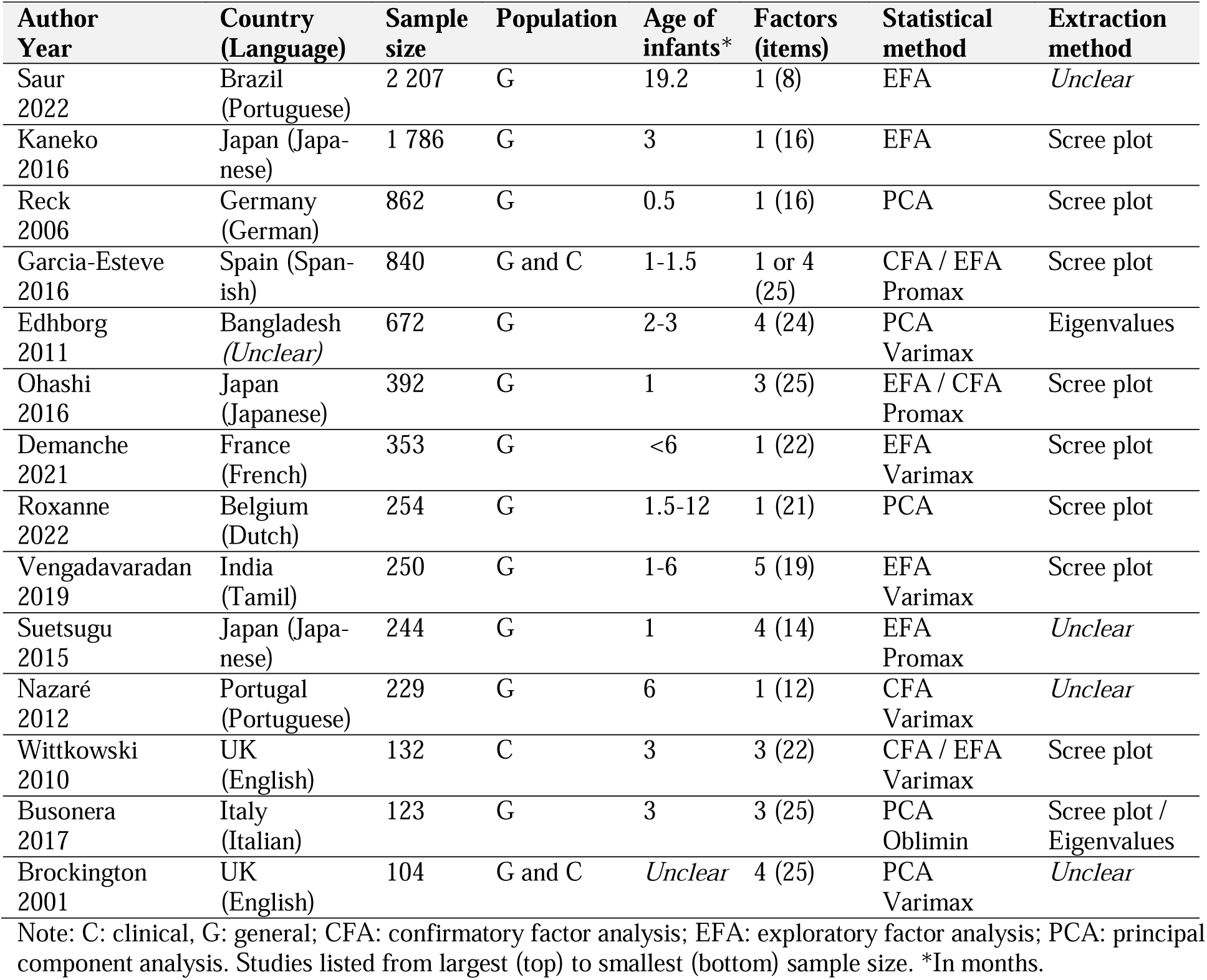
Characteristics of Psychometric Studies on the PBQ.

| Author<br>Year | Country<br>(Language) | Sample<br>size | Population | Age of<br>infants* | Factors<br>(items) | Statistical<br>method | Extraction<br>method |
| --- | --- | --- | --- | --- | --- | --- | --- |
| Saur<br>2022 | Brazil<br>(Portuguese) | 2 207 | G | 19.2 | 1 (8) | EFA | *Unclear* |
| Kaneko<br>2016 | Japan (Japanese) | 1 786 | G | 3 | 1 (16) | EFA | Scree plot |
| Reck<br>2006 | Germany<br>(German) | 862 | G | 0.5 | 1 (16) | PCA | Scree plot |
| Garcia-Esteve<br>2016 | Spain (Spanish) | 840 | G and C | 1-1.5 | 1 or 4<br>(25) | CFA / EFA<br>Promax | Scree plot |
| Edhborg<br>2011 | Bangladesh<br>(*Unclear*) | 672 | G | 2-3 | 4 (24) | PCA<br>Varimax | Eigenvalues |
| Ohashi<br>2016 | Japan<br>(Japanese) | 392 | G | 1 | 3 (25) | EFA / CFA<br>Promax | Scree plot |
| Demanche<br>2021 | France<br>(French) | 353 | G | <6 | 1 (22) | EFA<br>Varimax | Scree plot |
| Roxanne<br>2022 | Belgium<br>(Dutch) | 254 | G | 1.5-12 | 1 (21) | PCA | Scree plot |
| Vengadavaradan<br>2019 | India<br>(Tamil) | 250 | G | 1-6 | 5 (19) | EFA<br>Varimax | Scree plot |
| Suetsugu<br>2015 | Japan (Japanese) | 244 | G | 1 | 4 (14) | EFA<br>Promax | *Unclear* |
| Nazaré<br>2012 | Portugal<br>(Portuguese) | 229 | G | 6 | 1 (12) | CFA<br>Varimax | *Unclear* |
| Wittkowski<br>2010 | UK<br>(English) | 132 | C | 3 | 3 (22) | CFA / EFA<br>Varimax | Scree plot |
| Busonera<br>2017 | Italy<br>(Italian) | 123 | G | 3 | 3 (25) | PCA<br>Oblimin | Scree plot /<br>Eigenvalues |
| Brockington<br>2001 | UK<br>(English) | 104 | G and C | *Unclear* | 4 (25) | PCA<br>Varimax | *Unclear* |

Note: C: clinical, G: general; CFA: confirmatory factor analysis; EFA: exploratory factor analysis; PCA: principal component analysis. Studies listed from largest (top) to smallest (bottom) sample size. \*In months.

Further, little work has examined the factor structure of the PBQ in an English-speaking population from the United States (US), with the exception of one study reporting on the development of a shortened 10-item version of the PBQ (S-PBQ).^35^ Thus, using data from a US-based cohort of mothers at 4 months postpartum, the aim of the present study is two-tiered: 1) examine whether the original 25-item/4-factor structure proposed by Brockington and colleagues^21^ can be replicated, and 2) explore the factor structure and psychometric properties of the PBQ.

## Methods

### Participants

This study is a secondary analysis of data obtained from the prospective COVID-19 Mother Baby Outcomes (COMBO) Initiative and the Epidemiology of Severe Acute Respiratory Syndrome Coronavirus 2 in Pregnancy and Infancy (ESPI) COMBO (ESPI COMBO) sub-study. As part of these two parallel studies, we enrolled mother-infant dyads prenatally or up to 4 months postpartum and followed the health and wellbeing of mothers and infants through both cross-sectional and longitudinal study designs. Mother-infant dyads were enrolled beginning in March 2020 until August 2022 from Columbia University Irving Medical Center (CUIMC) in New York, New York for COMBO, and from CUIMC, University of Alabama in Birmingham, Alabama (UAB), and University of Utah (UU) in Salt Lake City, Utah for ESPI COMBO. Study procedures were approved by the CUIMC Institutional Review Board and written informed consent was obtained from all participants.

### Instruments

At 4 months postpartum, mothers completed the original 25-item PBQ in English via electronic self-administration (REDCap). Consistent with the original PBQ administration, items were scored on a 6-point Likert scale (0-Always to 5-Never). As described above, the PBQ comprises four subscales, i.e., general bonding disorders (12 items; 1, 2, 6, 7, 8, 9, 10, 12, 13, 15, 16, 17), severe mother-infant relationship disorders (7 items; 3, 4, 5, 11, 14, 21, 23), infant-focused anxiety (4 items; 19, 20, 22, 25), and risk of abuse (2 items; 18, 24).^21,34^ The total cumulative score, ranging from 0 to 125, is also used to screen for general bonding disorders (cut-off score ≥26), and severe bonding disturbances (cut-off score ≥40).

At 4 months postpartum, mothers also completed the Patient Health Questionnaire-9 (PHQ-9),^36^ one of the most reliable depression screening questionnaires.^37^ The PHQ-9 is a short 9-item self-administered questionnaire scored on a 4-point Likert scale (0-Not at all to 3-Nearly every day). The cumulative total score serves as an indicator of depression severity, ranging from no depression (scores 0-4), mild depressive symptoms (scores 5-9), moderate depressive symptoms (scores 10-14), moderately-severe depressive symptoms (scores 15-19), and severe depressive symptom (scores 20-27).^36^

### Statistical Analyses

Statistical analyses were conducted using SPSS v24 and R using the lavaan package.^38^ We first examined the demographic characteristics of the sample and the scale scores using descriptive statistics. For a comprehensive description of the PBQ factor scores and to screen for normality of the distributions, we examined medians/interquartile ranges, means/standard deviations as well and kurtosis and skewness.^39^ Before conducting the factorial analyses described below, consistent with the original scoring, 17 of the 25 PBQ items were reverse coded (2, 3, 5, 6, 7, 10, 12, 13, 14, 15, 17, 18, 19, 20, 21, 23 and 24), such that higher scores indicated lower bonding.

We tested the 4-factor structure proposed by Brockington and colleagues^21,34^ using confirmatory factor analysis (CFA) with maximum likelihood estimation. Global fit indices, such as comparative fit index (CFI), Tucker-Lewis index (TLI), root mean square error of approximation (RMSEA), relative chi-square (_χ_2/df), and item loadings were considered when evaluating model fit.

The factorial structure of the PBQ in our sample was investigated using a series of exploratory factor analyses (EFA) and the principal axis factoring extraction method. Bartlett’s test of sphericity and the Kaiser-Meyer-Olkin (KMO) measure of sampling adequacy were used to determine suitability of data for factorializability.^39^ The number of factors to retain was based on scree plot inspection and parallel analysis.^40^ Promax rotation was used to increase interpretability of the factor matrices.^41^ Items with factor loadings ≥.4 were considered meaningful and were therefore retained.^41^ The internal consistency of the scale and its factors were assessed using McDonald’s Omega (ω) coefficients.^42^ Omega coefficients ≤.69 were considered unacceptable, coefficients ranging from .70 to .79 were considered acceptable, from .80 to .89 were considered good, and >.90 were considered excellent.^43^

Finally, to further investigate the validity of the scale, we explored the association using Pearson correlations between our new PBQ and the original PBQ total score. Additionally, we explored the correlation (Pearson) between the new PBQ and the PHQ-9 scores, as previous psychometric studies have shown an association between higher depression scores and higher PBQ scores, such that higher maternal depression is associated with lower bonding.^22,23,25-28,30,32,33,44^

## Results

### Sample Characteristics

At 4 months postpartum, 610 mothers across CUIMC (n=331, 54.3%), UU (n=207, 33.9%), and UAB (n=72, 11.8%) completed the PBQ. Most mothers identified as White (n=368, 66.5%), Black (n=71, 12.8%), or Asian (n=32, 5.8%), and identified their ethnicity as not of Hispanic, Latinx, or Spanish origin (n=404, 72.7%). Mean age at survey completion was 32±5 years. A majority of mothers were married (n=246, 83.7%) and reported having completed some college education (n=444, 79.7%). Approximately half of the sample (n=234; 45.6%) was composed of first-time mothers. The mean score on the PHQ-9 was 3.43±3.72, with 69.3% (n=420) of mothers not meeting any cut-off for depression, 23.5% (n=142) meeting the cut-off for mild depression, 5.9% (n=36) for moderate depression, 0.5% (n=3) for moderately severe depression, and 0.8% (n=5) for severe depression. The mean infant age at the time of survey completion was 4.92±1.69 months, and 47.5% (n=288) of infants were female.

### Distribution of the Original PBQ Structure

The mean score on the 25-item PBQ was 9.38 (SD=7.17), and the mean scores on the four original factors were 5.21±3.85 on bonding disorders (factor 1), 1.78±2.40 on relationship disorders (factor 2), 2.38±1.99 on infant-focused anxiety (factor 3), and .01±.13 on risk of abuse (factor 4). The median skewness for all unique items was 2.5, thus the distribution of subscale data did not follow a Gaussian distribution (see Table 2). Based on conventional criteria, the internal consistency was poor for factor 3 (ω=.49), and not even estimable for factor 4.

**Table 2.** PBQ-25 distribution in the COMBO/ESPI COMBO cohort.

|  | Possible range* (cut-off<sup>†</sup>) | Median (IQR) | Mean (SD) | Meeting cut-off n(%) | Skewness (SE) | Kurtosis (SE) | McDonald's Omega |
| --- | --- | --- | --- | --- | --- | --- | --- |
| Factor 1 | 0-60 (12) | 5 (2-8) | 5.21 (3.85) | 44 (7.2%) | 0.99 (.10) | 1.78 (.20) | .74 |
| Factor 2 | 0-35 (13) | 1 (0-3) | 1.78 (2.40) | 2 (.3%) | 1.85 (.10) | 3.87 (.20) | .78 |
| Factor 3 | 0-20 (10) | 2 (1-4) | 2.38 (1.99) | 2 (.3%) | .69 (.10) | .13 (.20) | .54 |
| Factor 4 | 0-10 (3) | 0 (0-0) | .01 (.13) | 0 (0%) | 10.83 (.10) | 130.63 (20) | NA |
| Total | 0-125 (26) | 7 (4-12) | 9.38 (7.17) | 14 (2.3%) | 1.16 (.10) | 1.79 (.20) | .84 |

Note: NA=Not Applicable; McDonald's Omega cannot be estimated on less than 3 items.
\*Higher score indicates lower bonding.
<sup>†</sup>Scores above cut-off indicate high-risk for impairments.

### Confirmatory Factor Analysis

We first tried to replicate the 25-item/4-factor structure initially proposed by Brockington and colleagues.^21,34^ Although the model converged, no optimal solution was found and fit indices were not stable for interpretation, indicating poor fit of this factorial structure in our data.^45^

### Exploratory Factor Analysis

Given our failed attempt at confirming the original 4-factor structure, we examined the structure of the PBQ in our sample using a series of EFA. After removing item 18 (‘*I have done harmful things to my baby’*) because of low correlations (<.60) in the anti-image correlation matrix, all assumptions for EFA were met (Bartlett’s test: p<.001; KMO=.89). The number of factors to be retained was estimated by scree plot inspection and parallel analysis. Although 7 factors had eigenvalues>1, there was a sharp drop in scree plot slope after the first factor (Figure 1). In contrast, the parallel analysis supported a 3-factor structure. Therefore, both 1- and 3-factor solutions were further explored.

**Figure1.**
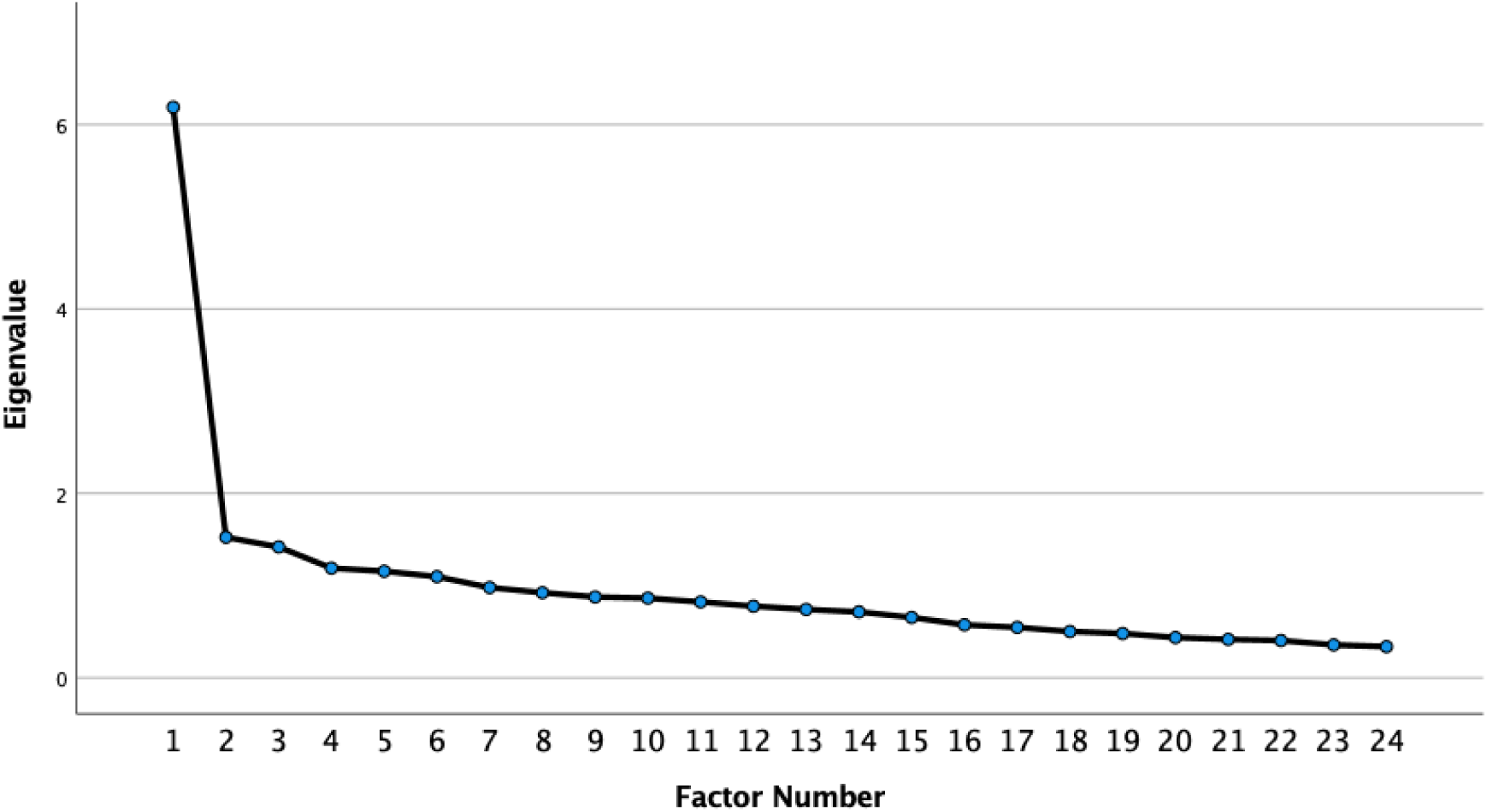
Scree plot.

The 1-factor solution accounted for 24.76% of the total variance and was characterized by general disruptions in mother-to-infant bond. After dropping items with loadings <.4, 14 items were retained (1, 2, 3, 4, 5, 10, 11, 12, 13, 14, 19, 21, 22, 23; see Table 3). The distribution of the new 14-item PBQ, hereafter referred to as PBQ-14, did not follow a Gaussian distribution (see Table 4), but internal consistency was high (ω=.87).

On the other hand, the 3 factors from the 3-factor configuration respectively explained 24.76%, 30.86% and 36.56% of the variance. After Promax rotation, 15 items with meaningful loadings, i.e., ≥.4 (see Table 3), were retained (hereafter referred to as PBQ-15). The first factor encompassed five items (1, 3, 4, 11, 16) which represented feelings of connection, e.g., ‘*I feel close to my baby’*, ‘*I love to cuddle my baby’*. The second factor included six items (10, 12, 14, 19, 21, 25) which, in contrast with the first factor, represented feelings of disconnection, e.g., ‘*My baby irritates me’*, ‘*My baby annoys me’*. Finally, four items relating to maternal rejection were retained on the third factor (5, 15, 17, 24), e.g., ‘*I resent my baby’*, ‘*I wish my baby would somehow go away’*. The three factors did not follow a Gaussian distribution (see Table 4), in addition to having unacceptable to low internal consistency (ω≤ .48 to .76). The inter-factor correlations were in the expected range and varied from *r=*.36 to *r*=.51 (p<.001).

**Table 3.** Factor Loadings.

|  |  | 1-Factor solution* | 3-Factor solution<sup>†</sup> |  |  |
| --- | --- | --- | --- | --- | --- |
|  |  | **Factor 1**<br>*General bonding*<br>*disruptions* | **Factor 1**<br>*Feelings of*<br>*connection* | **Factor 2**<br>*Feelings of dis-*<br>*connection* | **Factor 3**<br>*Feelings of*<br>*rejection* |
| 1 | I feel close to my baby | **.630** | **.763** | -.023 | -.039 |
| 2 | I wish the old days when I had no baby<br>would come back | **.564** | .159 | .192 | .292 |
| 3 | I feel distant from my baby | **.635** | **.576** | .015 | .119 |
| 4 | I love to cuddle my baby | **.580** | **.823** | .046 | -.228 |
| 5 | I regret having this baby | **.478** | .305 | -.184 | **.434** |
| 6 | The baby doesn't seem to be mine | .311 | .127 | -.108 | .345 |
| 7 | My baby winds me up | .161 | -.178 | .229 | .142 |
| 8 | I love my baby to bits | .210 | .178 | -.053 | .111 |
| 9 | I feel happy when my baby smiles or<br>laughs | .327 | .396 | -.053 | .018 |
| 10 | My baby irritates me | **.673** | .041 | **.686** | .042 |
| 11 | I enjoy playing with my baby | **.625** | **.682** | .031 | -.019 |
| 12 | My baby cries too much | **.434** | -.064 | **.760** | -.202 |
| 13 | I feel trapped as a mother | **.628** | .280 | .243 | .181 |
| 14 | I feel angry with my baby | **.597** | -.076 | **.486** | .370 |
| 15 | I resent my baby | .334 | -.048 | -.043 | **.496** |
| 16 | My baby is the most beautiful baby in<br>the world | .244 | **.477** | -.110 | -.107 |
| 17 | I wish my baby would somehow go<br>away | .364 | -.003 | -.214 | **.671** |
| 18<sup>‡</sup> | I have done harmful things to my baby | -- | -- | -- | -- |
| 19 | My baby makes me feel anxious | **.612** | .010 | **.486** | .205 |
| 20 | I am afraid of my baby | .286 | -.072 | .137 | .272 |
| 21 | My baby annoys me | **.698** | .142 | **.445** | .205 |
| 22 | I feel confident when caring for my<br>baby | **.428** | .164 | .123 | .197 |
| 23 | I feel the only solution is for someone<br>else to look after my baby | **.440** | .169 | .058 | .276 |
| 24 | I feel like hurting my baby | .197 | -.186 | -.021 | **.459** |
| 25 | My baby is easily comforted | .331 | -.017 | **.644** | -.258 |

Note: meaningful loadings (>.40) are bolded.
\*Loadings extracted with principal axis factoring.
<sup>†</sup>Loadings extracted with principal axis factoring and Promax rotation.
<sup>‡</sup>Item 18 was removed from the pool of items prior to running factorial analyses.

**Table 4.** Distribution of the 1-Factor and 3-Factor Solutions.

|  | Possible range* | Median (IQR) | Mean (SD) | Skewness (SE) | Kurtosis (SE) | McDonald's Omega |
| --- | --- | --- | --- | --- | --- | --- |
| **1-Factor solution** |  |  |  |  |  |  |
| Total | 0-70 | 5 (2-10) | 6.47 (5.65) | 1.17 (.10) | 1.43 (.20) | .87 |
| **3-Factor solution** |  |  |  |  |  |  |
| Factor 1 | 0-25 | 1 (0-2) | 1.40 (1.92) | 1.75 (.10) | 3.53 (.20) | .76 |
| Factor 2 | 0-30 | 3 (1-6) | 3.94 (3.12) | .85 (.10) | .42 (.20) | .74 |
| Factor 3 | 0-20 | 0 (0-0) | .23 (.73) | 4.34 (.10) | 22.16 (.20) | .48 |
| Total | 0-75 | 4 (2-8) | 5.57 (4.78) | 1.15 (.10) | 1.43 (.20) | .82 |

\*Higher score indicates lower bonding.

### Associations Between Original PBQ, New PBQ, and Maternal Depression

As expected, the correlations between the original PBQ and the new PBQ-14 (*r*=.96, p<.001), as well as the new PBQ-15 (*r*=.95, p<.001) were high, as was the correlation between the PBQ-14 and the PBQ-15 (*r*=.94, p<.001). Finally, we explored the associations between the new 1-factor PBQ-14 and the new 3-factor PBQ-15 with maternal depression. As expected, the PBQ-14 (*r*=.44, p<.001), as well as with the PBQ-15 factor scores (factor 1: *r*=.41, p<.001; factor 2: *r*=.36, p<.001; factor 3: *r*=.21, p<.001), all correlated significantly with PHQ-9 scores; thus associating maternal depression with bonding disruptions (PBQ-14), and with fewer feelings of connection (PBQ-15, factor 1), more feelings of disconnection (PBQ-15, factor 2), and more feelings of rejection (PBQ-15, factor 3).

## Discussion

In current pediatric clinical care, strong ERH is thought to be a crucial driver of optimal child-oriented life-course outcomes.^1,46^ In response to the 2021 American Academy of Pediatrics’ policy statement to universalize promotion of ERH,^1^ a systematic review has examined the global effectiveness of contemporary parent/caregiver-infant ERH interventions initiated within the first six months of life.^14^ Although significant improvements on several ERH outcomes were noted, meta-analytic results did not provide evidence of improved child socioemotional, mental, or behavioral health.^14^ These findings highlight the need for a more in-depth investigation of the mechanisms that are responsible for the emergence of strong ERH and later child positive outcomes. Given the importance of bonding as an early marker of ERH, addressing and communicating about measurement of parent/caregiver-to-infant bonding will add rigor to investigations of potential mechanisms driving positive ERH outcomes. Therefore, prior to undertaking mechanistic investigations of ERH, here we sought to confirm and explore the factorial structure of the PBQ as one of the most widely used measurements of parent/caregiver-reported bonding.

In alignment with other PBQ psychometric study results,^15,24,29,44^ the original 25-item/4-factor structure proposed by Brockington and colleagues^21,34^ could not be confirmed in our sample of English-speaking US-based mothers. Alternatively, following EFA, our data supported either a single dimension representing general bonding disruptions, or three factors differentiating bonding into parent/caregiver feelings of connection (factor 1), of disconnection (factor 2), and of rejection (factor 3).

Here, however, we argue that the added value of retaining structure beyond one factor seems modest and we suggest that a unidimensional scale is more suitable for the measurement of parent/caregiver-reported bonding. At the statistical level, only a minimal amount of total variance was explained by a second (6.1%) and a third factor (5.7%), in addition to having unacceptable to low internal consistency. From a conceptual perspective, bonding has consistently been described as encompassing the parent/caregiver’s feelings and emotions toward their infant.^16,18,19^ Although there was no evidence in our data of statistical redundancy among the three factors in the 3-factor configuration (PBQ-15), at the conceptual level, the more fine-grained differentiation between feelings of connection, disconnection and rejection doesn’t meaningfully add to the interpretation of bonding as described in the literature. In fact, it appears that items rather loaded together with regards to their valence despite all falling under the general umbrella of parent/caregiver-to-infant-directed feelings.

Thus, for practical purposes and conceptual coherence, we advocate for use of the 1-factor/14-item structure (PBQ-14). Although high variability in factor solutions (Table 1) and configurations of items retained (Figure 2) has been documented across languages, countries, and populations, other studies with adequate sample sizes (n>250) almost universally support a single factor solution to screen for general parent/caregiver-to-infant bonding disruptions. These striking differences in factor solutions may be a function of methodological variability and quality across studies, e.g., sample size, timing of assessment, statistical methods used, but they could also result from the lack of clear conceptual boundaries among different ERH outcomes or from the still poorly understood cultural influences in ERH perceptions and applications.

**Figure 2.**
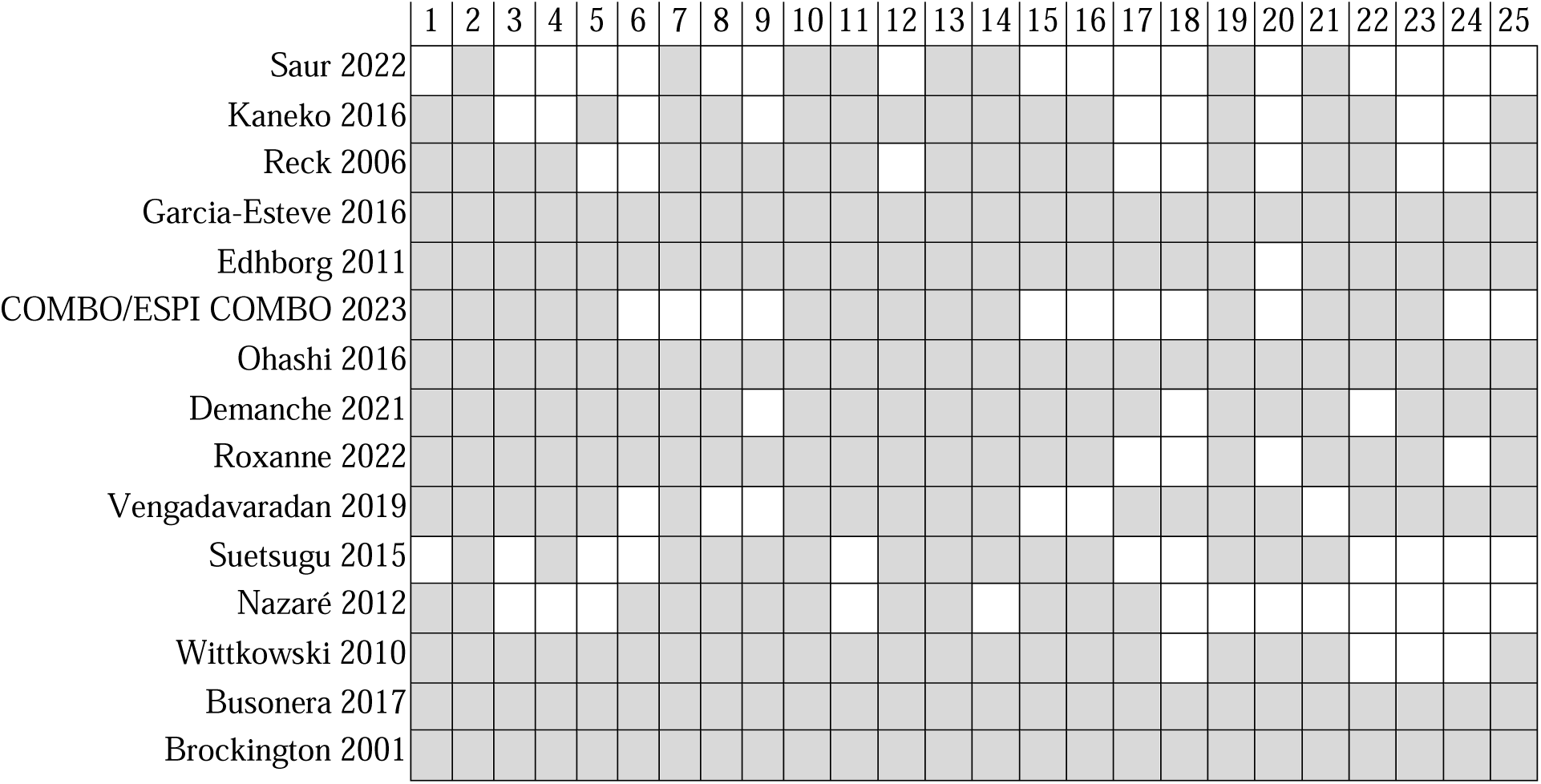
Overview of PBQ factor structures across psychometric studies. Distribution of the original 25 items included in factor structures across studies. Studies listed from largest (top) to smallest (bottom) sample size. Grey squares indicate retained items.

## Conclusion

The new unidimensional PBQ-14 has demonstrated good internal consistency and validity and is suitable for use in the US as a measure of general postnatal parent/caregiver-to-infant bonding disruptions. Future studies using the PBQ in the US should seek to confirm this new proposition using CFA. Nonetheless, as research efforts in the field of pediatrics are oriented toward a better understanding of the synergistic relationship between different aspects of ERH and child development and wellbeing, it is clear that a stringent scale development and validation agenda is needed for robust parent/caregiver-reported and observational measures of ERH.^15^

## Data Availability

The dataset analyzed here are available from the corresponding authors (AL or DD) upon reasonable request.

## Author Contributions

Substantial contributions to conception and design, acquisition of data, or analysis and interpretation of data: AL, JMW, MR, SL, JA, TB, MHK, MH, DD. Drafting the article or revising it critically for important intellectual content: AL, JMW, MR, SL, JA, MHK, TB, SE, DD. Final approval of the version to be published: AL, JMW, MR, SL, JA, MHK, TB, SE, MH, DD.

## Consent Statement

All study participants completed consent forms prior to participation following guidelines of the CUIMC Institutional Review Board.

